# Summaries, Analysis and Simulations of Recent COVID-19 Epidemic in Mainland China During December 31 2021-December 6 2022

**DOI:** 10.1101/2023.02.07.23285380

**Authors:** Lequan Min

## Abstract

**Background:** The recent COVID-19 epidemic in mainland China is an important issue for studying the prevention and disease control measures and the spread of the COVID-19 epidemic. Following our previous study for the mainland China epidemic during December 31 2021 to 30 April 2022, this paper studies and compares the the mainland China epidemic during December 31 2021 to December 6, 2022.

**Methods:** Using differential equations and real word data (both domestic and foreign input infected individuals) modelings and simulates COVID-19 epidemic in mainland China during May 1 2022 to December 6 2022, estimates the transmission rates, the recovery rates, the blocking rates to the symptomatic and the asymptomatic infections, and the died rate of the symptomatic infected individuals. The transmission rates and the recovery rates of the foreign input COVID-19 infected individuals in mainland China have also been studied. Using virtual simulations predict the outcomes of the epidemics.

**Results:** The simulation results were in good agreement with the real word data.

- The average input transmission rate of the foreign input symptomatic infection individuals was much lower than the average transmission rates of the symptomatic infection causing by the mainland symptomatic and asymptomatic individuals.
- The average input transmission rate of the foreign input asymptomatic infection individuals was was much lower than the average transmission rate of the asymptomatic infection causing by the mainland symptomatic individuals.
- The average recovery rates of the foreign input COVID-19 symptomatic and asymptomatic infected individuals were much higher than the average recovery rates of the mainland symptomatic and asymptomatic infected individuals.

For the mainland epidemic simulations:

- If kept the transmission rates, the recovery rates, the death rate and the blocking rates on day 181 (June 30, 2022), the numbers of the current symptomatic and asymptomatic individuals would reduce to about one on day 270 (September 27, 2022).
- If kept the transmission rates, the recovery rates, the death rate and the blocking rates on day 340 (December 6, 2022) until day 380 (January 15, 2023), the numbers of the current symptomatic and the asymptomatic infected individuals would increase to 37 999 and 224 945, respectively, the cumulative death individuals would increase from 599 to 616.
- If kept the transmission rates, the recovery rates on day 340, but decreased the blocking rates to 34% and select the death rate to equal to the average death rate during days 104-150, then the simulation showed that on day 380, the numbers of current symptomatic and the asymptomatic infected individuals would increase to about 323 559 095 and 481 270 717, respectively, and the cumulative death individuals would reach about 1 055 607.

For the foreign input epidemic simulations:

- If kept the transmission rates, the recovery rates, and the blocking rates day 242 (August 30, 2022), until day 340, the numbers of the current symptomatic and the asymptomatic infected individuals would decrease to 13 and 430, respectively.
- If kept the transmission rates, the recovery rates, and the blocking rates on day 340 until day 380 (January 15, 2023), the numbers of the current symptomatic and the asymptomatic infected individuals would decrease and increase to 168 and 1952, respectively.
- Recommendations on COVID-19 epidemic base on WHO’s technic guidelines and HBV infection experiment in Chimpanzees are provided.

**Conclusions:**

- For the mainland individuals’ epidemic, keeping the blocking rates of over 86% and 93% to the symptomatic and asymptomatic infections, and the recovery rates of over 0.119 and 0.112 to the symptomatic and asymptomatic individuals may make the numbers of the current symptomatic and asymptomatic infected individuals to decrease to very low levels in three months.
- For the foreign input individuals’ epidemic, keeping the transmission rates of under 0.07 to the symptomatic and asymptomatic infections, and the recovery rates of over 0.125 and 0.099 to the symptomatic and asymptomatic individuals may make the numbers of the current symptomatic and asymptomatic infected individuals to decrease to very low levels in four months.
- After December 6 2022, decreasing the blocking rates of under 34% to the symptomatic and asymptomatic infections may cause over 1100 millions individuals’ COVID-19 infections and over one million COVID-19 infected individuals’ death.
- It is necessary that administrations implement strict prevent and control strategies to prevent the spread of new COVID-19 variants.

## 1 Introduction

Globally, nearly 1.9 million new COVID-19 cases and over 12 000 deaths were reported in the week of 16 to 22 January 2023. In the last 28 days (26 December 2022 to 22 January 2023), over 11 million cases and over 55 000 new deaths were reported globally [1].

Mainland China prevents effectively the spread of COVID-19 epidemics before 2022. Omicron and Delta variant virus appearing makes the numbers of the symptomatic and the asymptomatic COVID-19 infected individuals to increase rapidly.

In a previous paper [2](also see [3]), we have studied the mainland China epidemic during December 31 2021 to 30 April 2021. This paper summarizes, analyzes and simulates the recent COVID-19 epidemic in mainland China (December 31 2021 - December 6 2022), estimates the infection transmission rates, the infection blocking rates, and the preventive measures through modelings and numerical simulations.

## 2 Materials and Methods

The dataset of the China COVID-19 epidemics from December 31, 2021 to December 6, 2022 was collected and edited from the National Health Commission of the People’s Republic of China official website [4]. Using differential equation models [2,3,5] stimulates the outcomes of the numbers of the current symptomatic individuals, the current asymptomatic individuals charged in medical observations, the cumulative recovered symptomatic individuals and cumulative asymptomatic individuals discharged from medical observations. Equation parameters were determined by so-called minimization error square criterion described in references [2, 3, 5]. Using virtual simulations estimates outcomes of the spreads of COVID-19 epidemics in mainland China. Simulations and figure drawings were implemented via Matlab programs.

## 3 Analysis and Simulations of COVID-19 Epidemics in China

### 3.1 COVID-19 Epidemics in Mainland

Figure 1 shows the outcomes of the numbers of the current symptomatic individuals (CSI) charged in observation and the current asymptomatic individuals (CAI) charged in observations. Figure 2 shows the outcomes of the numbers of the cumulative recovered symptomatic individuals (CCSI) and the cumulative asymptomatic individuals (CCAI) discharged from observations.

**Figure 1:**
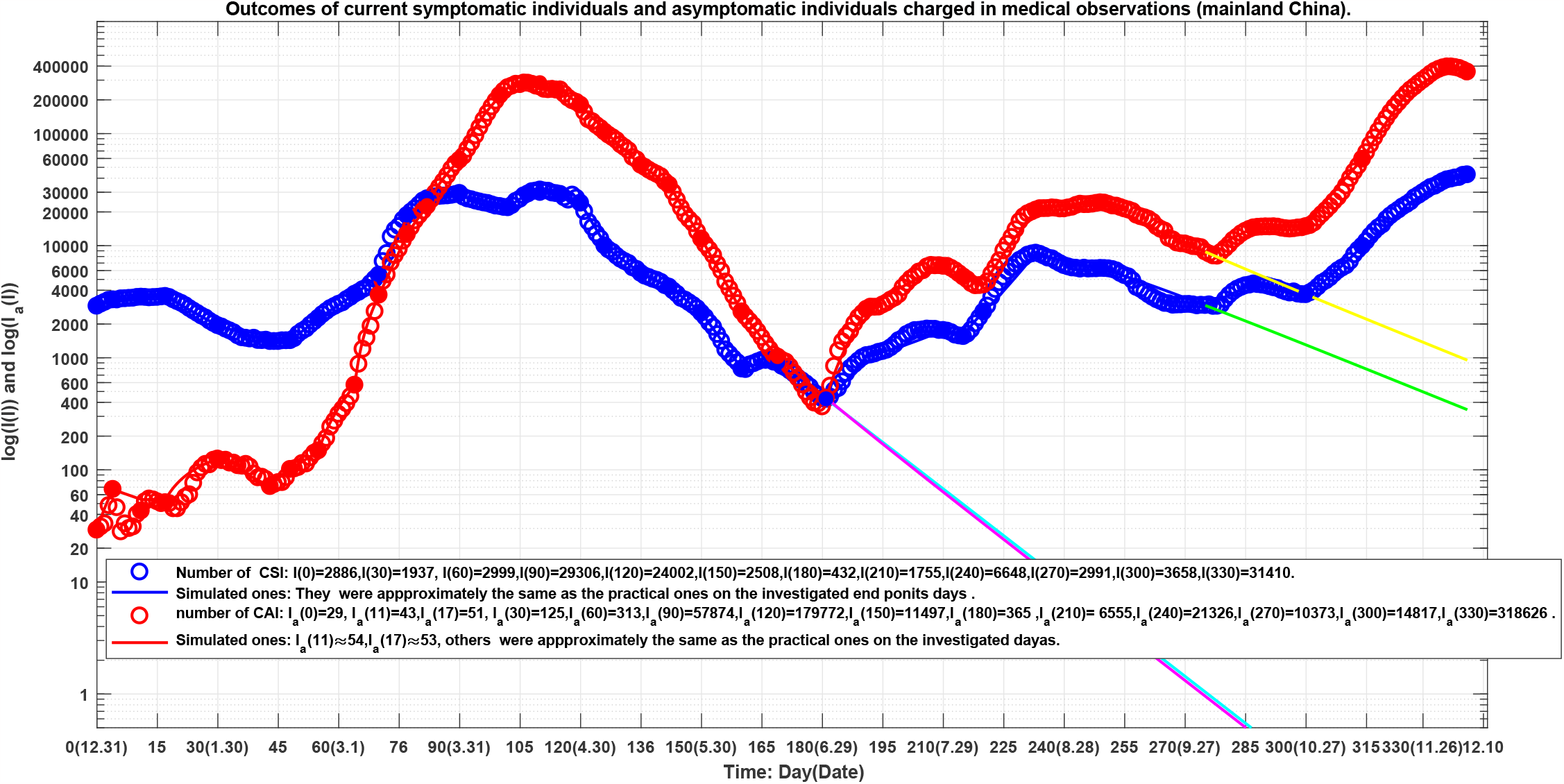
Blue circles: outcome of the number of the current symptomatic individuals (CSI), blue line: outcome of the corresponding simulations of equation (1). Red circles: outcome of the numbers of the current asymptomatic individuals (CAI) charged in medical observations, red line: outcome of the corresponding simulations of equation (1). The lines colored by cyan, magenta, green and yellow correspond to the virtual simulation results of equation (1). See Section Mainland Epidemic Virtual Simulations for details.

**Figure 2:**
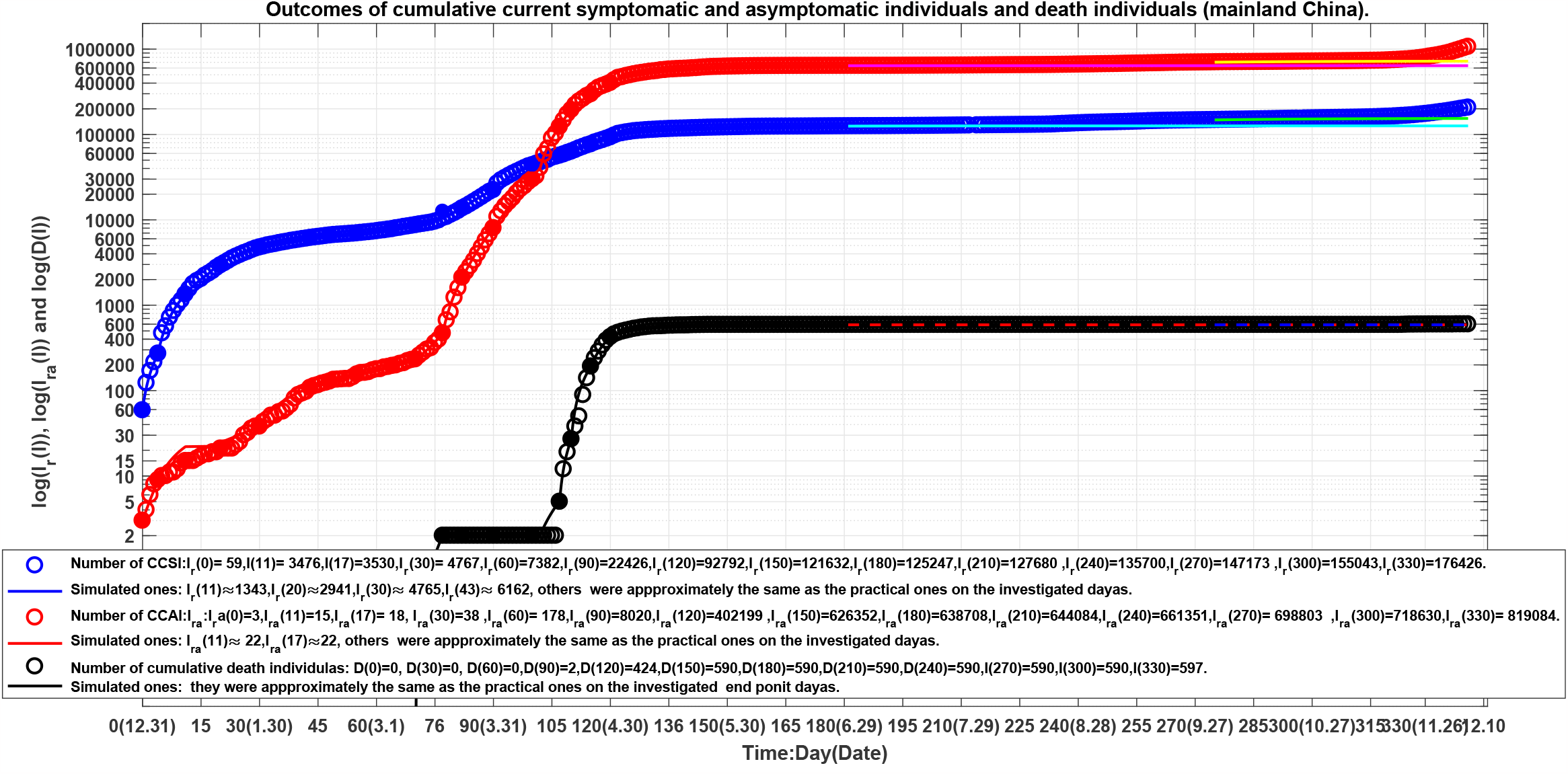
Blue circles: the outcome of the number of the current cumulative recovered symptomatic individuals (CCSI), blue line: outcome of the corresponding simulations of equation (1). Red circles: the outcome of the numbers of the current cumulative asymptomatic individuals (CCAI) discharged in medical observations, red line: outcome of the corresponding simulations of equation (1). The lines colored by cyan, magenta, green and yellow correspond to the virtual simulation results of equation (1). See Section Mainland Epidemic Virtual Simulations for details.

The recent COVID-19 epidemics in mainland China are still continuing. Although there existed several turning points and valley points of the current symptomatic infections, for examples on days 17 (January 17), 43 (February 11), 90 (March 31), 110 (April 20), 181 (June 30), 233 (August 11), and 300 (October 27). The COVID-19 epidemics made that the China authority declared the “open prevent control” policy on December 7, 2022.

In order to estimate numerically the transmission rates and the blocking rates to the symptomatic and the asymptomatic infections, we need to set up mathematics models (similar to [2, 3, 5, 6]) to simulate the dynamics of the spread of the infection disease.

For the mainland epidemics over the *lth* transmission interval, the current symptomatic infected individuals (*I*) and the current asymptomatic infected individuals (*I*_*a*_) infect the susceptible population (*S*) with the transmission rates of *β*_11_(*l*) and *β*_21_(*l*), respectively, making *S* become symptomatic infected individuals, and with the transmission rates of *β*_12_(*l*) and *β*_22_(*l*), respectively, making *S* become asymptomatic individuals. Then, a symptomatic individual is cured at a rate *κ*(*l*), an asymptomatic individual returns to normal at a rate *κ*_*a*_(*l*). Here all parameters are positive numbers. Assume that the dynamics of an epidemic can be described by *m*-time intervals, which correspond different transmission rates, prevention and control measures, and medical effects. At *l*th time interval, the model has the form described by equation (1) in [2, 3, 5, 6]). Where Θ_1_(*i*) = (1 − *θ*_1_(*l*)) and Θ_2_(*l*) = (1 − *θ*_2_(*l*)) (*l* = 1, …, *m*) represent the blocking rates to the symptomatic and asymptomatic infections, respectively.

It can be assumed that the input transmissions can be divided into 37 time intervals (see solid points in Figs. 1 and 2). We need to determine the parameters of equation (1) for *l* = 1, 2, …, 37. Over the *lth* time interval [*t*_*l*−1_, *t*_*l*_], denote

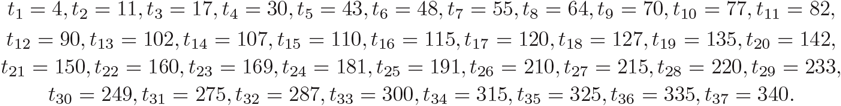

Using the minimization error square criterion ([2, 3, 5]) determines the *β*_*ij*_(*l*)′*s, κ*(*l*)′*s, κ*_*a*_(*l*)′*s*), *θ*_1_(*l*)′*s*, and *θ*_2_(*l*)′*s*. The calculated parameters are shown in Table 1. The corresponding simulation results of the equation (see (1) given in [2, 3, 5]) are shown in Fig. 1 and Fig. 2.

**Table 1:**
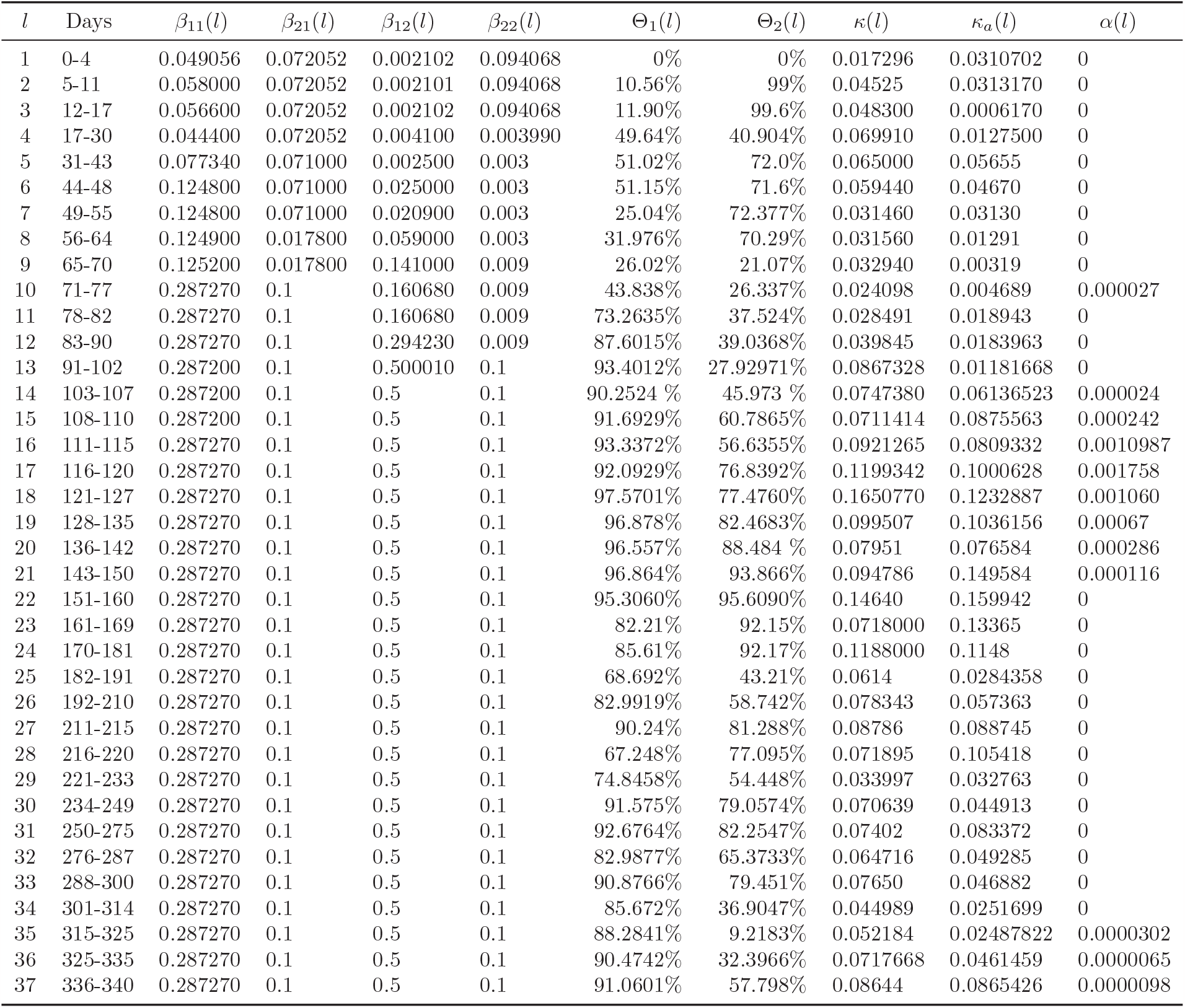
The equation parameters of the COVID-19 epidemics in mainland China during December 1 2021 - December 6 2022.

Observe that the simulation results of the equation were in good agreement with the data of the COVID-19 epidemics in mainland China (see Table 2, and the solid blue lines, the red lines in Fig. 1 and Fig. 2).

**Table 2:** Data set of the COVID-19 epidemics in mainland China on the investigated point days.

| Day | Data | $I(l)$ | $I_a(l)$ | $I_r(l)$ | $I_{ra}(l)$ | $D(l)$ |
| --- | --- | --- | --- | --- | --- | --- |
| 0 | 2021.12.31 | 2886 | 29 | 59 | 3 | 0 |
| 4 | 2022. 01.04 | 3291 | 67 | 272 | 9 | 0 |
| 11 | 2022. 01.11 | 3476 | 43 | 1344 | 15 | 0 |
| 17 | 2022. 01.17 | 3530 | 51 | 2359 | 18 | 0 |
| 30 | 2022. 01.30 | 1937 | 125 | 4767 | 38 | 0 |
| 43 | 2022. 02.11 | 1397 | 71 | 6164 | 108 | 0 |
| 48 | 2022. 02.17 | 1423 | 101 | 6583 | 128 | 0 |
| 55 | 2022. 02.24 | 2254 | 148 | 6981 | 155 | 0 |
| 64 | 2022. 03.04 | 3691 | 570 | 7808 | 195 | 0 |
| 70 | 2022. 03.13 | 5461 | 3624 | 8698 | 233 | 0 |
| 77 | 2022. 03.18 | 18586 | 12597 | 10505 | 470 | 2 |
| 82 | 2022. 03.23 | 26253 | 22564 | 13661 | 2110 | 2 |
| 90 | 2022. 03.31 | 29306 | 57874 | 22426 | 8020 | 2 |
| 102 | 2022. 04.12 | 21826 | 257493 | 47165 | 40654 | 2 |
| 107 | 2022. 04.17 | 28987 | 281992 | 56674 | 123194 | 5 |
| 110 | 2022. 04.20 | 31421 | 260517 | 63136 | 194345 | 27 |
| 115 | 2022. 04.25 | 28726 | 245543 | 76970 | 296741 | 192 |
| 120 | 2022. 04.30 | 24002 | 179772 | 92792 | 402199 | 424 |
| 127 | 2022. 05.07 | 9181 | 97007 | 110537 | 518277 | 538 |
| 135 | 2022. 05.15 | 5661 | 52320 | 116348 | 578226 | 577 |
| 142 | 2022. 05.22 | 4275 | 34763 | 119101 | 601237 | 587 |
| 150 | 2022. 05.30 | 2508 | 11497 | 121632 | 626352 | 590 |
| 160 | 2022. 06.09 | 795 | 2571 | 123826 | 635868 | 590 |
| 169 | 2022. 06.18 | 897 | 1014 | 124387 | 637848 | 590 |
| 181 | 2022. 06.30 | 426 | 436 | 125290 | 638788 | 590 |
| 191 | 2022. 07.10 | 1047 | 2712 | 125698 | 639180 | 590 |
| 210 | 2022. 07.29 | 1755 | 6555 | 127680 | 644084 | 590 |
| 215 | 2022. 08.03 | 1548 | 5268 | 128406 | 646694 | 590 |
| 220 | 2022. 08.08 | 2561 | 4468 | 129145 | 649220 | 590 |
| 233 | 2022. 08.11 | 8583 | 20791 | 131309 | 653891 | 590 |
| 249 | 2022. 09.06 | 6280 | 24163 | 139508 | 670346 | 590 |
| 275 | 2022. 10.02 | 2929 | 8814 | 148152 | 702888 | 590 |
| 287 | 2022. 10.14 | 4563 | 14442 | 151007 | 709647 | 590 |
| 300 | 2022. 10.27 | 3658 | 14817 | 155043 | 718630 | 590 |
| 314 | 2022. 11.10 | 9915 | 59141 | 158879 | 730195 | 590 |
| 325 | 2022. 11.21 | 23296 | 226934 | 167527 | 765140 | 595 |
| 335 | 2022. 12.01 | 38748 | 394333 | 189250 | 905111 | 597 |
| 340 | 2022. 12.06 | 42955 | 354890 | 207015 | 1066737 | 599 |

### 3.2 Results and Discussions

Recent China COVID-19 epidemics with both Omicron and Delta variants make more difficult to prevent spread of the diseases. On the end points of the 37 investigated time-intervals [*t*_*l*−1_, *t*_*l*_]′*s*, that is on day 0, day 4, day 11, day 17, day 30, day 43, day 48, day 55, day 64, day 70 day 77, day 82, day 90, day 102, day 107, day 110, day 115, day 120, day 127, day 135, day 142, day 150, day 160, day 169, day 181, day 191, day 210, day 215, day 220, day 233, day 249, day 275, day 287, day 300, day 325, day 325, day 335, and day 340, we obtain the following results (also see Figs. 1 and 2 and Table 2).

1. On the end point days of the 37 investigated time-intervals, the numbers of the reported and the simulated current symptomatic individuals were approximate the same (errors were less than one individual).
2. On day 11 and day 17, there were eleven and four differences between the numbers of the reported and the simulated current asymptomatic individuals. On the other end point days, the numbers of the reported and the simulated current asymptomatic individuals charged in medical observations were approximate the same.
3. On the end point days of the 37 investigated time-intervals, the numbers of the reported and the simulated current cumulative recovered symptomatic individuals were approximate the same (errors were less than one individual).
4. On day 11 and day 17, there were seven and four differences between the numbers of the reported and the simulated current cumulative asymptomatic individuals discharged from observations. On the other end point days, the numbers of the reported and the simulated current cumulative asymptomatic individuals discharged from medical observations were approximate the same.
5. On the end point days of the 37 investigated time-intervals, the numbers of the reported cumulative died symptomatic individuals and the simulated ones were approximate the same (errors were less than one individual).

- Generally speaking, the transmission rates *β*_*ij*_ of the infections increased during the investigated days. During the first 170 days, the maximal 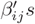 *s* were

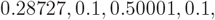

respectively. The minimal 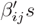 *s* were

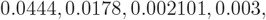

respectively. The average transmission rates were about

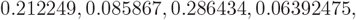

respectively. During the last 170 days, the transmission rates *β*_*ij*_ un-changed, they were

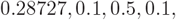

respectively.
- The recovery rates *κ*(*l*)′*s* of the symptomatic infection individuals and the asymptomatic infection individuals waved. During the first 170 days, the maximal recovery rates of the symptomatic infection and asymptomatic infection reached to 0.165077 and 0.159942, respectively. The minimal recovery rates of the symptomatic infection and asymptomatic infection reached to 0.017296, and 0.000617, respectively. The average recovery rates of were about 0.071423, and 0.061583, respectively. During the last 170 days, the maximal recovery rates of the symptomatic infected individuals and the asymptomatic infected individuals reached to 0.118 and 0.1148, respectively. The minimal recovery rates of the symptomatic infected individuals and asymptomatic infected individuals reached to 0.033997 and 0.024878, respectively. The average recovery rates were about 0.071017 and 0.059622, respectively.
- The transmission rates of the symptomatic infections caused by the symptomatic individuals were increasing from day 31 to day 77, and then seems to stop (see *β*_11_(*l*)′*s* in Table 1).
- The transmission rates of the asymptomatic infections caused by the symptomatic individuals have obviously increased from day 44 to day 102, and seems to stop until the investigated end point day (see *β*_12_(*l*)′*s* in Table 1).
- During days 0-55, the transmission rate of the symptomatic infections caused by the asymptomatic individuals has kept stable after two weeks’ decreasing rapidly, the transmission rate increased to 0.1 and then seems stop until the investigated end point day (see *β*_21_(*l*)′*s* in Table 1).
- The transmission rates of the asymptomatic infections caused by the asymptomatic individuals obviously increased after day 90 and kept at very low level 0.1 until the investigated end point day (see *β*_22_(*l*)′*s* in Table 1).
- The blocking rates Θ_1_(*l*)′*s* and Θ_2_(*l*)′*s* to symptomatic and asymptomatic infections were not hight. Even on day 181 the valley day (see Fig. 1 and Table 2), the blocking rates only reached to about 85.61% and 92.17.0% (see Tables 1 and 2), respectively. However, for the first and second epidemics in Beijing and the five wave epidemics in Shanghai, the blocking rates reached to over 95% in one month [3, 5, 6].
- The recovery rates *κ*(*l*) and *κ*_*a*_(*l*), and the blocking rates Θ_1_(*l*)′*s* and Θ_2_(*l*)′*s* of the symptomatic individuals and the asymptomatic individuals waved.
- During days 0-43, the recovery rates increased which made the numbers of the symptomatic individuals and the asymptomatic individuals decreased from local maximal 3291 and 125 to local minimum 1397 and 71, respectively (see Fig. 1, Tables 1 and 2).
- During days 44-77, The blocking rates Θ_1_(*l*)′*s* and Θ_2_(*l*)′*s* and the recovery rates *κ*(*l*) and *κ*_*a*_(*l*) decreased in wave ways, which made the numbers of the symptomatic individuals and the asymptomatic individuals decreased from local minimum 1397 and 71, to local maximum 29306 and 57874 on day 90, respectively (see Fig. 1, Tables 1 and 2).
- During days 77 - 181, The blocking rates Θ_1_(*l*)′*s* and Θ_2_(*l*)′*s*, and the recovery rates *κ*(*l*) and *κ*_*a*_(*l*) decreased in wave ways, which made the numbers of the symptomatic individuals and the asymptomatic individuals decreased from local maximum 29306 and 57874, to local minimum 426 and 436, respectively (see Fig. 1, Tables 1 and 2).
- During days 182-233, the blocking rates and the recovery rates to the infections decreased significantly in wave ways, which made the numbers of the symptomatic individuals and the asymptomatic individuals increased from 426 and 436 to 8583 and 20791, respectively (see Fig. 1, Tables 1 and 2).
- During days 234-300, the blocking rates and the recovery rates to the infections increased significantly in wave ways, which made the numbers of the symptomatic individuals and the asymptomatic individuals decreased from 8583 and 20791 to 3658 and 14817, respectively (see Fig. 1, Tables 1 and 2).
- During days 301-335, the blocking rates and the recovery rates to the asymptomatic infections decreased significantly in wave ways, which made the numbers of the symptomatic individuals and the asymptomatic individuals increased from 3658 and 14817 to 38748 and 394333, respectively (see Fig. 1, Tables 1 and 2).
- During days 335-340, the blocking rates and the recovery rates to the asymptomatic infections increased significantly, which made the numbers of the symptomatic individuals and the asymptomatic individuals increased and decreased from 38748 and 394333 to 42955 and 354890, respectively (see Fig. 1, Tables 1 and 2).
- During days 0-103, there were only two death cases appearing. However during days 104-150, the death cases increased to 590, which happen in Shanghai, and the average death rate was about 0.0006563375. During days 315-340, there were new nine death cases appearing, and the average death rate was about 0.000015733 (see Table 1).

### 3.3 Foreign input COVID-19 Epidemics in China

Figure 3 shows the outcomes of the numbers of the current symptomatic individuals (CSI) and the current asymptomatic individuals (CAI). Figure 4 shows the outcomes of the numbers of the cumulative recovered symptomatic individuals (CCSI) and the cumulative asymptomatic individuals (CCAI) discharged from observations.

**Figure 3:**
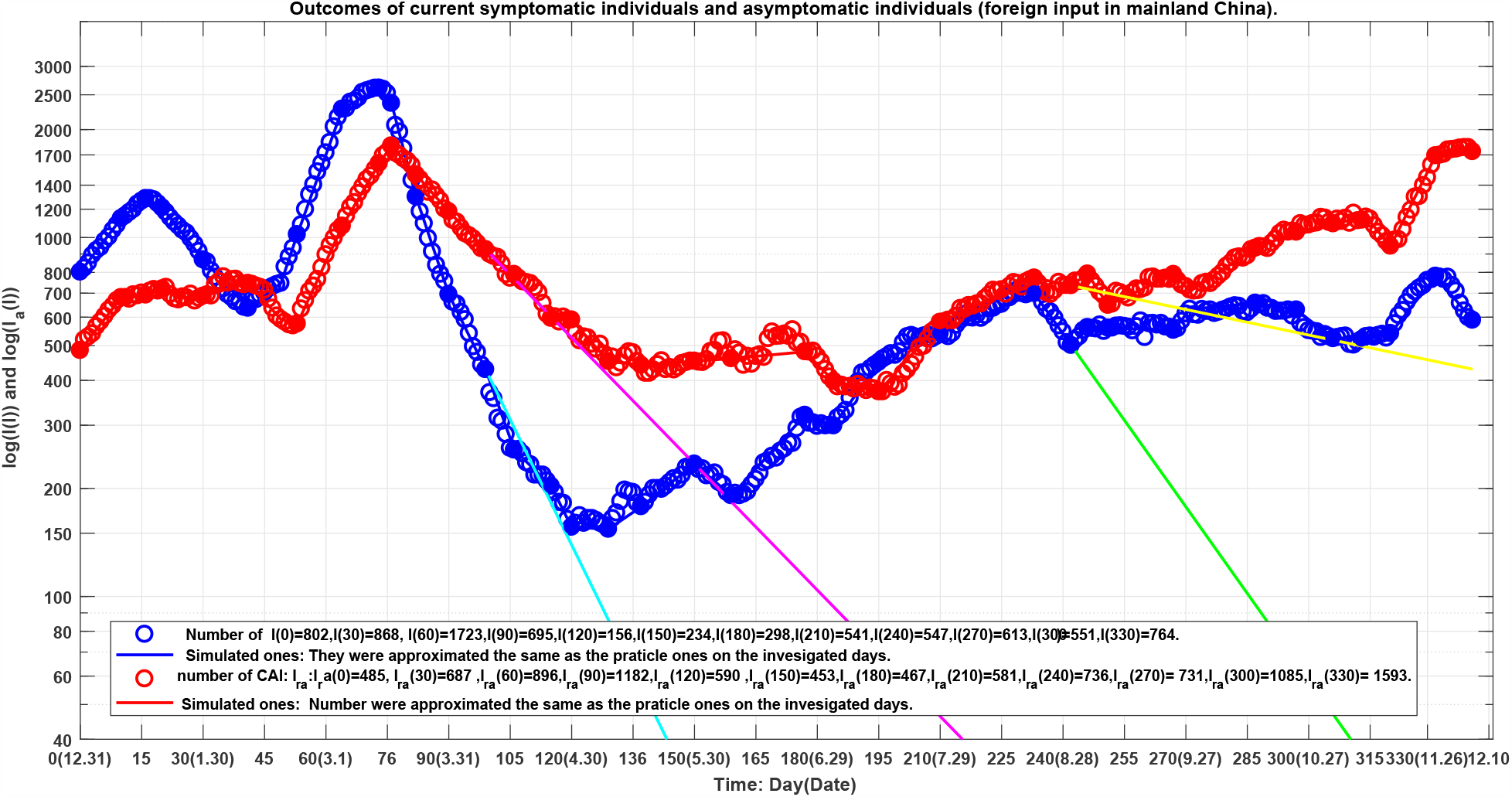
Blue circles: outcome of the number of the current symptomatic individuals (CSI), blue line: outcome of the corresponding simulations of equation (2). Red circles: outcome of the numbers of the current asymptomatic individuals (CAI) charged in medical observations, red line: outcome of the corresponding simulations of equation (2). The lines colored by cyan and magenta correspond to the virtual simulation results of equation (2). See Section Foreign Input Epidemic Virtual Simulations for details.

**Figure 4:**
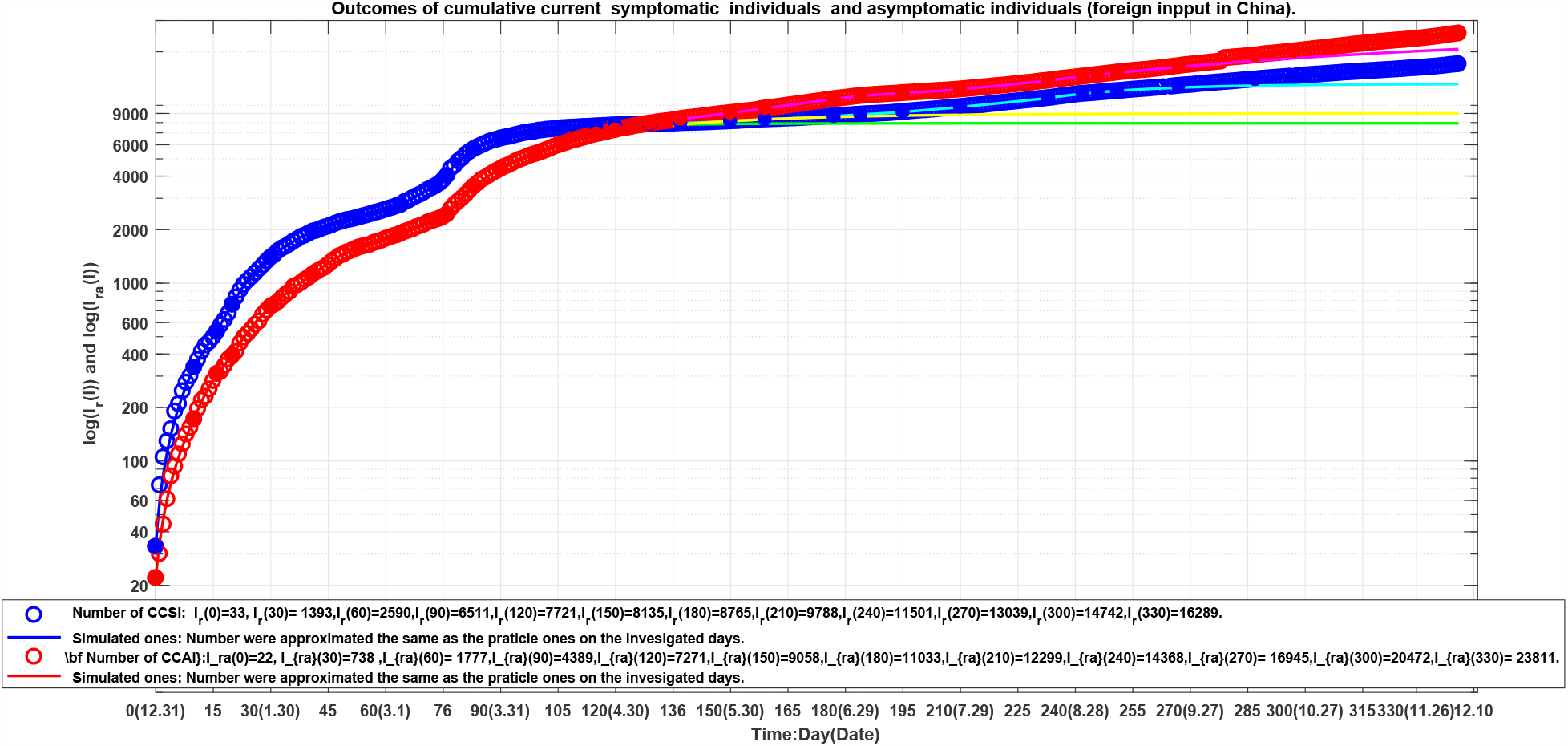
Blue circles: outcome of the number of the current cumulative recovered symptomatic individuals (CCSI), blue line: outcome of the corresponding simulations of equation (2). Red circles: outcome of the numbers of the current cumulative asymptomatic individuals (CCAI) discharged in medical observations, red line: outcome of the corresponding simulations of equation (2). The lines colored by cyan and magenta correspond to the virtual simulation results of equation (2). See Section Foreign Input Epidemic Virtual Simulations for details.

Generally speaking the foreign input epidemics were under control although the asymptomatic infection increased rapidly in the last month. There existed several turning points and valley points of the current symptomatic infections, for examples on days 15 (January 16), 41 (February 9), 73 (March 14), 129 (May 7), 181 (June 30), 233 (August 21), and 242 (August 30), 287 (October 14), 320 (November 16) (see Table 4).

For the foreign input COVID-19 infected individuals, they were discovered immediately and no further transmissions generated each other after entering China. Therefore the model has simply the form given in equation (2) in reference [2, 3] where *β*_11_(*l*) and *β*_22_(*l*) represent input transmission rates of the symptomatic individuals and asymptomatic individuals over the *lth* time interval, respectively.

It can be assumed that the input transmissions can be divided into 37 time intervals (see solid points in Fig. 3 and Fig. 4) We need to determine the parameters for equation (2) for *l* = 1, 2, …, 37 over the *lth* time interval [*t*_*l*−1_, *t*_*l*_]. Denote

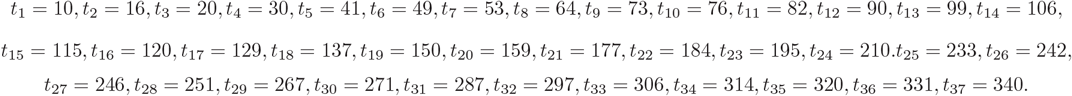

Using the minimization error square criterion [2, 3] determines the *β*_11_(*l*)′*s, β*_22_(*l*)′*s, κ*(*l*)′*s* and *κ*_*a*_(*l*)′*s*. The calculated parameters are shown in Table 3. The corresponding simulation results of equation (2) are shown in Fig. 3 and Fig. 4. Observe that the simulation results of equation (2) were in good agreement with the data of the foreign input COVID-19 epidemics (see Table 4 and the solid blue lines and the red lines in Fig. 3 and Fig. 4). On the end points of the 37 investigated time-interval [*t*_*l*−1_, *t*_*l*_]′*s*, the simulated numbers and the actual reported numbers were approximate the same–errors were less than one person, respectively.

**Table 3:** The equation parameters of the foreign input COVID-19 epidemics in mainland China during 2021.12.31-2022.5.15.

| $l$ | Days | $\beta_{11}(l)$ | $\beta_{22}(l)$ | $\kappa(l)$ | $\kappa_a(l)$ |
| --- | --- | --- | --- | --- | --- |
| 1 | 0-10 | 0.065980 | 0.059898 | 0.031686 | 0.025982 |
| 2 | 11-16 | 0.048874 | 0.036028 | 0.027328 | 0.033269 |
| 3 | 17-20 | 0.029875 | 0.038568 | 0.044693 | 0.028747 |
| 4 | 21-30 | 0.028319 | 0.044800 | 0.061721 | 0.049466 |
| 5 | 31-41 | 0.039880 | 0.055930 | 0.068146 | 0.048460 |
| 6 | 41-49 | 0.071617 | 0.033290 | 0.051650 | 0.062170 |
| 7 | 50-53 | 0.108100 | 0.051367 | 0.029960 | 0.058598 |
| 8 | 54-64 | 0.097320 | 0.092677 | 0.024133 | 0.035659 |
| 9 | 65-73 | 0.049520 | 0.075400 | 0.03446950 | 0.030710 |
| 10 | 74-76 | 0.034320 | 0.069930 | 0.066998 | 0.032389 |
| 11 | 77-82 | 0.038166 | 0.060467 | 0.138124 | 0.091284 |
| 12 | 83-90 | 0.053600 | 0.069155 | 0.131850 | 0.098770 |
| 13 | 91-99 | 0.070690 | 0.071710 | 0.124300 | 0.098700 |
| 14 | 100-106 | 0.060960 | 0.0984000 | 0.134700 | 0.121250 |
| 15 | 107-115 | 0.063640 | 0.105230 | 0.089380 | 0.136545 |
| 16 | 116-120 | 0.051440 | 0.125180 | 0.104307 | 0.127220 |
| 17 | 121-129 | 0.081280 | 0.108270 | 0.082600 | 0.13768 |
| 18 | 130-137 | 0.096950 | 0.12695 | 0.07900 | 0.13000 |
| 19 | 138-150 | 0.094100 | 0.11900 | 0.07300 | 0.01170 |
| 20 | 151-159 | 0.844000 | 0.13520 | 0.10700 | 0.13340 |
| 21 | 160-177 | 0.10473 | 0.149139 | 0.07605 | 0.14670 |
| 22 | 178-184 | 0.08320 | 0.120490 | 0.09280 | 0.147448 |
| 23 | 185-195 | 0.10930 | 0.095400 | 0.07240 | 0.101600 |
| 24 | 196-210 | 0.09510 | 0.110670 | 0.08266 | 0.080900 |
| 25 | 211-233 | 0.09644 | 0.108814 | 0.08530 | 0.09647 |
| 26 | 234-242 | 0.07978 | 0.105230 | 0.11680 | 0.11070 |
| 27 | 243-246 | 0.11682 | 0.123550 | 0.08810 | 0.104850 |
| 28 | 247-251 | 0.08200 | 0.088610 | 0.08270 | 0.128730 |
| 29 | 252-267 | 0.09476 | 0.126917 | 0.09566 | 0.114550 |
| 30 | 268-271 | 0.11400 | 0.110000 | 0.08050 | 0.137000 |
| 31 | 272-287 | 0.090601 | 0.174900 | 0.08809 | 0.157600 |
| 32 | 288-297 | 0.066700 | 0.115250 | 0.07108 | 0.104990 |
| 33 | 298-306 | 0.099480 | 0.115700 | 0.11996 | 0.109740 |
| 34 | 307-314 | 0.099700 | 0.110220 | 0.09800 | 0.104860 |
| 35 | 315-320 | 0.090300 | 0.079250 | 0.08690 | 0.11032 |
| 36 | 321-331 | 0.114850 | 0.131310 | 0.08148 | 0.07855 |
| 37 | 332-340 | 0.084370 | 0.098312 | 0.11570 | 0.09539 |

### 3.4 Results and Discussions

It seems that the foreign input COVID-19 infected individuals have been obtained good managements and therapies. On the end points of the 37 investigated time-interval [*t*_*l*−1_, *t*_*l*_]′*s*, that is on day 0, day 10, day 16, day 20, day 30, day 41, day 49, day 53, day 64, day 73, day 76, day 82, day 90, day 99, day 106, day 115, day 120, day 129, day 137, day 150, day 159, day 177, day 184, day 195, day 210, day 233, day 242, day 246, day 251, day 267,day 271, day 287, day 297, day 306, day 314, day 320, day 331 and day 340, we obtain the following results (also see Figs. 3 and 4).

- On the end points of the 37 investigated time-interval [*t*_*l*−1_, *t*_*l*_]′*s*, the numbers of the reported and the simulated current symptomatic individuals, the reported and the simulated current asymptomatic individuals charged in medical observations, the reported and the simulated current cumulative recovered symptomatic individuals, and the reported and the simulated current cumulative asymptomatic individuals discharged from medical observations were approximate the same.
- The foreign input transmission rates of the symptomatic infection individuals and the asymptomatic infection individuals waved. During the first 170 days, the maximal transmission rates of the symptomatic infection and asymptomatic infection reached to 0.1081 and 0.149139, respectively. The minimal transmission rates of the symptomatic infection and asymptomatic infection reached to 0.028319 and 0.03329, respectively. The average transmission rates were about 0.065417 and 0.082219, respectively. During the last 170 days, the maximal foreign input transmission rates of the symptomatic infection and asymptomatic infection reached to 0.11682 and 0.1749, respectively. The minimal input transmission rates of the symptomatic infection and asymptomatic infection reached to 0.0667 and 0.07925, respectively. The average transmission rates were about 0.095419 and 0.115515, respectively.
- The recovery rates *κ*(*l*)′*s* and *κ*_*a*_(*l*)′*s* of the foreign input symptomatic infection individuals and the asymptomatic infection individuals waved. During the first 170 days, the maximal recovery rates of the symptomatic infection and asymptomatic infection reached to 0.138124 and 0.1467, respectively. The minimal recovery rates of the symptomatic infection and asymptomatic infection reached to 0.024133, and 0.025982, respectively. The average recovery rates of were about 0.075290, and 0.083056, respectively.
- During the last 170 days, the maximal recovery rates of the symptomatic infection and asymptomatic infection reached to 0.1188 and 0.13365, respectively. The minimal recovery rates of the symptomatic infection and asymptomatic infection reached to 0.033995 and 0.024878, respectively. The average recovery rates were about 0.090246 and 0.113553, respectively.
- During days 0-16, the transmission rates *β*_11_(*t*), *β*_22_(*t*) and the recovery rate *κ*(*t*) decreased. However which still made the numbers of the current symptomatic individuals and the asymptomatic individuals to increase from 802 and 485 to local maximal values (turning points) 1286 and 720 on day 16 and day 20, respectively (see Fig.3 and Tables 3 and 4).

**Table 4:** The data set of the foreign input COVID-19 individuals in mainland China on the investigated point days.
- During days 17-41, the transmission rates *β*_11_(*t*), *β*_22_(*t*) and the recovery rate *κ*(*t*) decreased and increased in wave ways, which made the numbers of the current symptomatic individuals and the asymptomatic individuals to decrease form 1286 and 720 to local minimal values (valley points) 636 and 575 on day 41 and day 53, respectively (see Fig.3 and Tables 3 and 4).
- During days 42-73, the transmission rates *β*_11_(*t*), *β*_22_(*t*) and the recovery rate *κ*(*t*) increased and decreased in wave ways, which made the numbers of the current symptomatic individuals and the asymptomatic individuals to increase from 636 and 575 to local maximal values 2612 and 1802 on day 73 and day 76, respectively (see Fig.3 and Tables 3 and 4).
- During days 74-129, the transmission rates *β*_11_(*t*), *β*_22_(*t*) and the recovery rate *κ*(*t*) decreased and increased in wave ways, which made the numbers of the current symptomatic individuals and the asymptomatic individuals to decrease form 2612 and 1802 to local minimal values 154 and 441 on day 129 and day 137, respectively (see Fig.3 and Tables 3 and 4).
- During days 130-233, the transmission rates *β*_11_(*t*), *β*_22_(*t*) and the recovery rate *κ*(*t*) increased and decreased in wave ways, which made the numbers of the current symptomatic individuals and the asymptomatic individuals to increase from 154 and 441 to local maximal values 699 and 772 on day 233, respectively (see Fig.3 and Tables 3 and 4).
- During days 234-242, the transmission rates *β*_11_(*t*), *β*_22_(*t*) and the recovery rate *κ*(*t*) decreased and increased, which made the numbers of the current symptomatic individuals and the asymptomatic individuals to decrease from 699 and 772 to local minimal values 501 and 535 on day 242, respectively (see Fig.3 and Tables 3 and 4).
- During days 243-306, the transmission rates *β*_11_(*t*), *β*_22_(*t*) and the recovery rate *κ*(*t*) changed in wave ways, which made the number of the current symptomatic individuals and the asymptomatic individuals reach to local minimal values 523 and 1097 on days 306 and 310, respectively (see Fig.3 and Tables 3 and 4).
- During days 307-320, the transmission rates *β*_11_(*t*), *β*_22_(*t*) and the recovery rate *κ*(*t*) changed in wave ways, which made the numbers of the current symptomatic individuals individuals to reach local minimal values 525 and 947 on days 319 and 320, respectively (see Fig.3 and Tables 3 and 4).
- During days 321-331, the transmission rates *β*_11_(*t*), *β*_22_(*t*) and the recovery rate *κ*(*t*) increase and decrease, respectively, which made the numbers of the current symptomatic individuals individuals to reach local maximal value 781 and maximal 1692 on day 331, respectively (see Fig.3 and Tables 3 and 4).
- During days 332-340, the transmission rates *β*_11_(*t*), *β*_22_(*t*) and the recovery rate *κ*(*t*) decreased and increased, respectively,which made the numbers of the current symptomatic individuals and individuals to decrease from 781 and 1783 (on day 338) to 589 and 1737, respectively (see Fig.3 and Tables 3 and 4).

| Day | Date | $I(l)$ | $I_a(l)$ | $I_r(l)$ | $I_{ra}(l)$ |
| --- | --- | --- | --- | --- | --- |
| 0 | 12.31 | 802 | 485 | 33 | 22 |
| 1 | 1.01 | 822 | 519 | 73 | 30 |
| 10 | 1.10 | 1130 | 681 | 336 | 172 |
| 16 | 1.16 | 1286 | 692 | 534 | 309 |
| 20 | 1.20 | 1212 | 720 | 757 | 390 |
| 30 | 1.30 | 868 | 687 | 1393 | 738 |
| 41 | 2.10 | 636 | 746 | 1952 | 1120 |
| 53 | 2.22 | 1020 | 575 | 2342 | 1588 |
| 64 | 3.05 | 2281 | 1077 | 2758 | 1902 |
| 73 | 3.14 | 2612 | 1610 | 3516 | 2268 |
| 76 | 3.17 | 2368 | 1802 | 4016 | 2434 |
| 82 | 3.23 | 1300 | 1498 | 5492 | 3335 |
| 90 | 3.31 | 695 | 1182 | 6511 | 4389 |
| 99 | 4.09 | 429 | 927 | 7128 | 5321 |
| 106 | 4.16 | 256 | 790 | 7444 | 6048 |
| 115 | 4.25 | 203 | 596 | 7628 | 6894 |
| 120 | 4.30 | 156 | 590 | 7721 | 7271 |
| 129 | 5.09 | 154 | 452 | 7836 | 7914 |
| 137 | 5.17 | 178 | 441 | 7941 | 8378 |
| 150 | 5.30 | 234 | 453 | 8135 | 9058 |
| 159 | 6.08 | 191 | 460 | 8339 | 9606 |
| 177 | 6.26 | 320 | 481 | 8681 | 10848 |
| 184 | 7.03 | 299 | 398 | 8882 | 11300 |
| 195 | 7.14 | 449 | 372 | 9176 | 11730 |
| 210 | 7.29 | 541 | 581 | 9788 | 12299 |
| 233 | 8.21 | 699 | 772 | 10998 | 13790 |
| 242 | 8.30 | 501 | 735 | 11623 | 14541 |
| 246 | 9.03 | 562 | 792 | 11810 | 14861 |
| 251 | 9.08 | 560 | 648 | 12042 | 15323 |
| 271 | 9.28 | 632 | 715 | 13084 | 17050 |
| 267 | 9.24 | 552 | 790 | 12893 | 16636 |
| 287 | 10.14 | 657 | 935 | 13991 | 19106 |
| 297 | 10.24 | 629 | 1036 | 14448 | 20140 |
| 306 | 11.02 | 523 | 1093 | 15068 | 21191 |
| 314 | 11.10 | 530 | 1141 | 15481 | 22128 |
| 320 | 11.16 | 541 | 947 | 15760 | 22817 |
| 331 | 11.27 | 781 | 1692 | 16346 | 23926 |
| 340 | 12.06 | 589 | 1737 | 17055 | 25398 |

### 3.5 Comparing

- During the first 170 days, the average input transmission rate of the foreign input symptomatic infected individuals was about 0.065417, which was much lower than the average transmission rates 0.212249 and 0.084867 of the symptomatic infection causing by the mainland symptomatic and asymptomatic individuals (see *β*_11_(*l*)′*s, β*_21_(*l*)′*s* in Table 1 and *β*_11_(*l*)′*s*, in Table 3).
- During the last 170 days, the average input transmission rate of the foreign input symptomatic infected individuals was about 0.095419, which was much lower than the average transmission rates 0.28727 and 0.1 of the symptomatic infection causing by the mainland symptomatic and asymptomatic individuals (see *β*_11_(*l*)′*s, β*_21_(*l*)′*s* in Table 1 and Table *β*_11_(*l*)′*s*, in Table 3).
- During the first 170 days, the average input transmission rate of the foreign input asymptomatic infected individuals was about 0.08219, which was much lower and higher than the average transmission rates 0.255924 and 0.05877 of asymptomatic infection caused by the mainland symptomatic and asymptomatic individuals, respectively (see *β*_21_(*l*)′*s, β*_22_ in Table 1 and *β*_22_(*l*)′*s* in Table 3).
- During the last 170 days, the average input transmission rate of the foreign input asymptomatic infected individuals was about 0.115515, which was much lower and slight higher than the average transmission rates 0.5 and 0.1 of the asymptomatic infection caused by the mainland symptomatic and the symptomatic individuals, respectively (see *β*_21_(*l*)′*s* in Table 1 and *β*_22_(*l*)′*s* in Table 3).
- During the first 170 days, the average recovery rates of the foreign input COVID-19 symptomatic and asymptomatic infected individuals were about 0.075290 and 0.083056, respectively, which were much higher the average recovery rates 0.065578 and 0.05093 of the mainland COVID-19 symptomatic and asymptomatic infected individuals, respectively (see *κ*(*l*)′*sκ*_*a*_(*l*)′*s* in Table 1 and Table 3).
- During last 170 days, the average recovery rates of the foreign input COVID-19 symptomatic and asymptomatic infected individuals were about 0.090246 and 0.113552, respectively, which were much higher than the average recovery rates 0.076895 and 0.075170 of the mainland COVID-19 symptomatic and asymptomatic infected individuals, respectively (see *κ*(*l*)′*sκ*_*a*_(*l*)′*s* in Table 1 and Table 3).
- During December 31, 2021 to December 6, 2022, the numbers of the symptomatic (hospitalized) infected individuals and the asymptomatic (charged in medical observations) infected individuals in mainland China increased from 2886 and 29 to 42955 and 354890, respectively. The numbers of the correspond foreign input individuals changed form 802 and 485 to 589 and 1737, respectively.

In summary,

1. the average input transmission rate of the foreign input symptomatic infection individuals was much lower than the average transmission rates of the symptomatic infection causing by the mainland symptomatic and asymptomatic individuals;
2. the average input transmission rate of the foreign input asymptomatic infection individuals was was much lower than the average transmission rate of the asymptomatic infection causing by the mainland symptomatic individuals;
3. The average recovery rates of the foreign input COVID-19 symptomatic and asymptomatic infected individuals were much higher than the average recovery rates of the mainland COVID-19 symptomatic and asymptomatic infected individuals, respectively.
4. By December 6, 2022, the outcomes of the foreign input epidemics were controlled; the outcomes of the mainland epidemics caused a surge.

## 4 Virtual Simulations

### 4.1 Mainland Epidemic Virtual Simulations

1. Assume that after day 181 (June 30, 2022), it still keeps the transmission rates 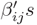 *s*, the blocking rates (Θ_1_(24), Θ_2_(24)), the recovery rates (*κ*(24), *κ*_*a*_(24)), and the death rate *α*(24) until day 340 (December 6, 2022). The simulation results of equation (1) are shown in Fig. 1 and Fig. 2 by cyan lines and magenta lines, respectively. Calculated results show that on day 277 (October 4, 2022), the numbers of the current symptomatic and the current asymptomatic infected individuals would decrease to about one, the numbers of the cumulative recovered symptomatic and asymptomatic individuals, and the cumulative died individuals would reach about 126086, 639536, and 590, respectively. On day 340, the numbers of current symptomatic and the asymptomatic infected individuals, the cumulative recovered symptomatic individuals, and the cumulative asymptomatic and individuals and the cumulative died individuals would reach about 0, 0, 126087, 639538, and 590, respectively.
2. Assume that after day 275 (October 2, 2022), it still keeps the transmission rates *β*_*ij*_(31)′*s*, the blocking rates (Θ_1_(31), Θ_2_(31)), the recovery rates (*κ*(1(31)), *κ*_*a*_(31)), and the death rate *α*(31) until day 340. The simulation results of equation are shown in Fig. 1 and Fig. 2 by green lines and yellow lines, respectively. Calculated results show that on day 340, the numbers of the current symptomatic and the current asymptomatic infected individuals would reach about 344 and 952, respectively. The numbers of the cumulative recovered symptomatic individuals and the cumulative asymptomatic individuals would reach about 154062 and 721794, and the cumulative died individuals would reach about 590, respectively.
3. Assume that after day 340 (December 6, 2022), it still keeps the transmission rates *β*_*ij*_(37)′*s*, the blocking rates (Θ_1_(37), Θ_2_(37)), the recovery rates (*κ*(1(37)), *κ*_*a*_(37)), and the death rate *α*(37) until day 380 (January 15, 2023). The simulation results show that on day 380, the numbers of the current symptomatic and the asymptomatic infected individuals would increase to 37 999 and 224 945, respectively. The numbers of cumulative recovered symptomatic individuals and cumulative asymptomatic individuals discharged in medical observations would reach about 354 787 and 2 024 416 and the cumulative death individuals would reach about 616, respectively.
4. Assume that after day 340 (December 6, 2022), it still keeps the transmission rates *β*_*ij*_(37)′*s*, the recovery rates *κ*(1(37)), *κ*_*a*_(37), and reduce the the blocking rates to (Θ_1_, Θ_2_) = (34%, 34%), select death rate *α* = 0.0006563375 (the average death rate during days 104-150) until day 380 (January 15, 2023). The simulation results show that on day 380, the numbers of current symptomatic and the asymptomatic infected individuals would increase to about 323 559 095 and 481 270 717, respectively. The numbers of cumulative recovered symptomatic individuals and cumulative asymptomatic individuals discharged in medical observations would reach about 139 046 349 and 208 034 530, and the cumulative death individuals would reach about 1 055 607, respectively.

### 4.2 Foreign Input Epidemic Virtual Simulations

1. Assume that after day 99 (April 9, 2022), it still keeps the transmission rates *β*_11_(13), *β*_22_(13), the recovery rates *κ*(13), and *κ*_*a*_(13) until day 340 (December 6, 2022). The simulation results of equation (2) are shown in Fig. 3 and Fig. 4 by cyan lines and magenta lines, respectively. Calculated results show that on day 215 (August 3), the numbers of the current symptomatic and the current asymptomatic infected individuals would reduce to about 1 and 41, respectively; On day 340 the numbers of the asymptomatic infected individuals cumulative recovered symptomatic individuals and the cumulative asymptomatic individuals discharged from medical observations would reach about 0, 1, 8123, and 8706, respectively.
2. Assume that after day 242 (August 30, 2022), it still keeps the transmission rates *β*_11_(26), *β*_22_(26), the recovery rates *κ*(26), and *κ*_*a*_(26) until day 340. The simulation results of equation (2) are shown in Fig. 3 and Fig. 4 by green lines and yellow lines, respectively. Calculated results show that on day 340, the numbers of the current symptomatic and the current asymptomatic infected individuals would reduce to about 13 and 430, respectively; the numbers of the cumulative recovered symptomatic individuals and the cumulative recovered asymptomatic individuals would reach about 13162 and 20712, respectively.

However if increased the recover rates to *κ* = *κ*(11) =0.138124, and *κ*_*a*_ = *κ*(31) =0.1576, then on day 340, the numbers of the symptomatic and asymptomatic infected individuals, the cumulative recovered symptomatic individuals and the cumulative recovered asymptomatic individuals would decrease to 1, 3, 12650, and 16105, respectively.

## 5 Recommendations on COVID-19 Epidemic based on HBV Infection in Chimpanzees and WHO’s technic guidelines

Chronic hepatitis B virus (HBV) infection is a major global health problem. HBV is the most serious type of viral hepatitis. About two billion people worldwide have been infected with the HBV. More than 350 million people have chronic (long-term) liver infection [7].

In the experiments on acute hepatitis B virus (HBV) infection in chimpanzees, Asabe et al. examined the impact of size of the viral inoculum on the outcome of HBV infection in nine healthy, native, immunocompetent adult chimpanzees by using a wide doses range (1 ∼ 10^1^0 copies). They observed that both high dose (10^10^ GE) and low dose (10^0^ GE) inocula primed the CD4 T cell response, allowing infection of 100% of hepatocytes and requiring prolonged immunopathology before clearance occurred [8].

Figure 5 shows the course of acute HBV infection of the chimpanzees after experimental inoculation with 1GN/animal ∼ 10^10^GN/animal of HBV. Figure 6 shows the outcomes of HBV DNA and ALT in chimpanzee A006 with 10^10^ GN/animal and chimpanzee Ch1618 with 1GN/animal.

**Figure 5:**
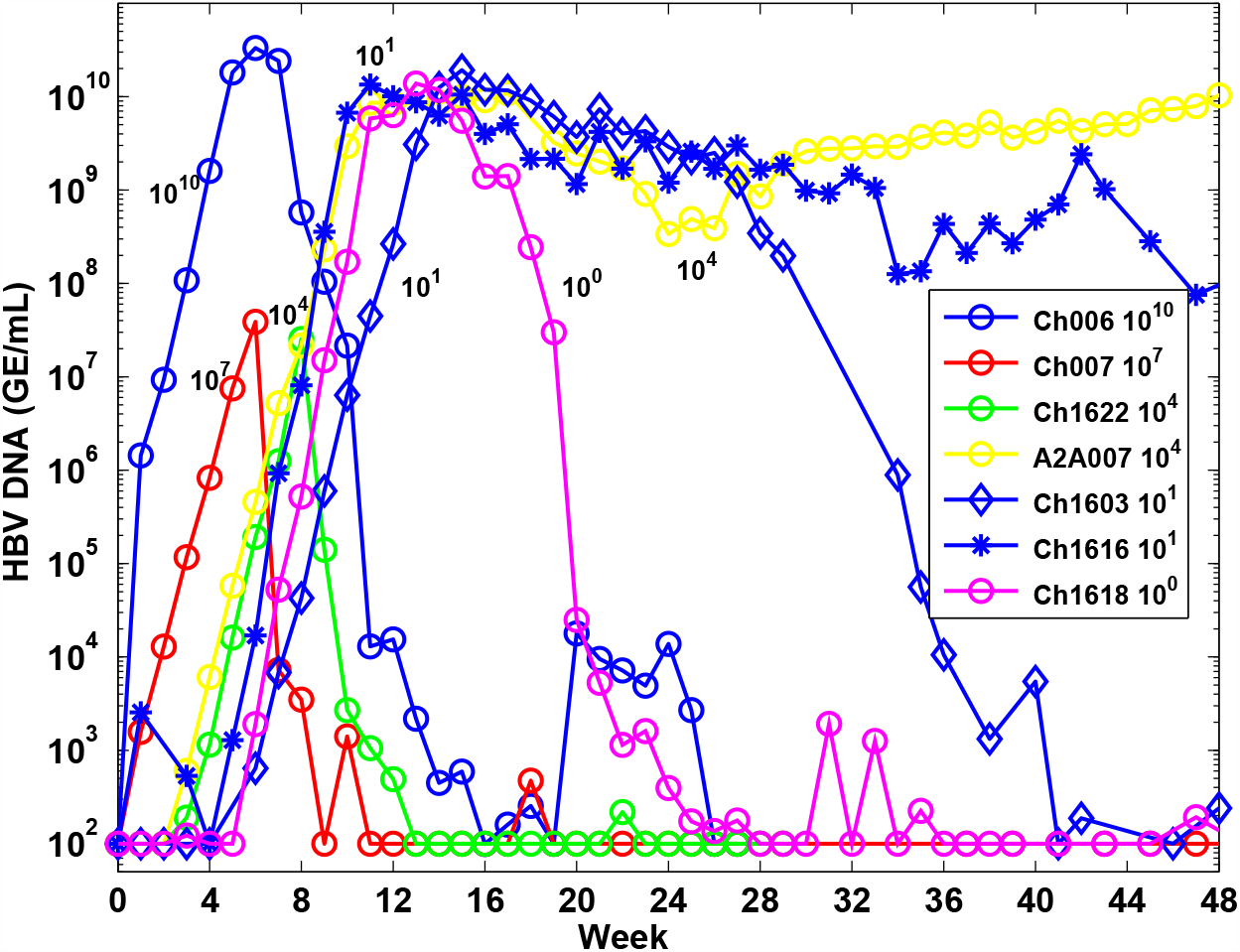
The outcomes of HBV DNA in chimpanzees with 1GN/animal ∼ 10^10^GN/animal (Thank Dr. S. F. Weiland at The Scripps Research Institute for proving the experimental data of the chimpanzees).

**Figure 6:**
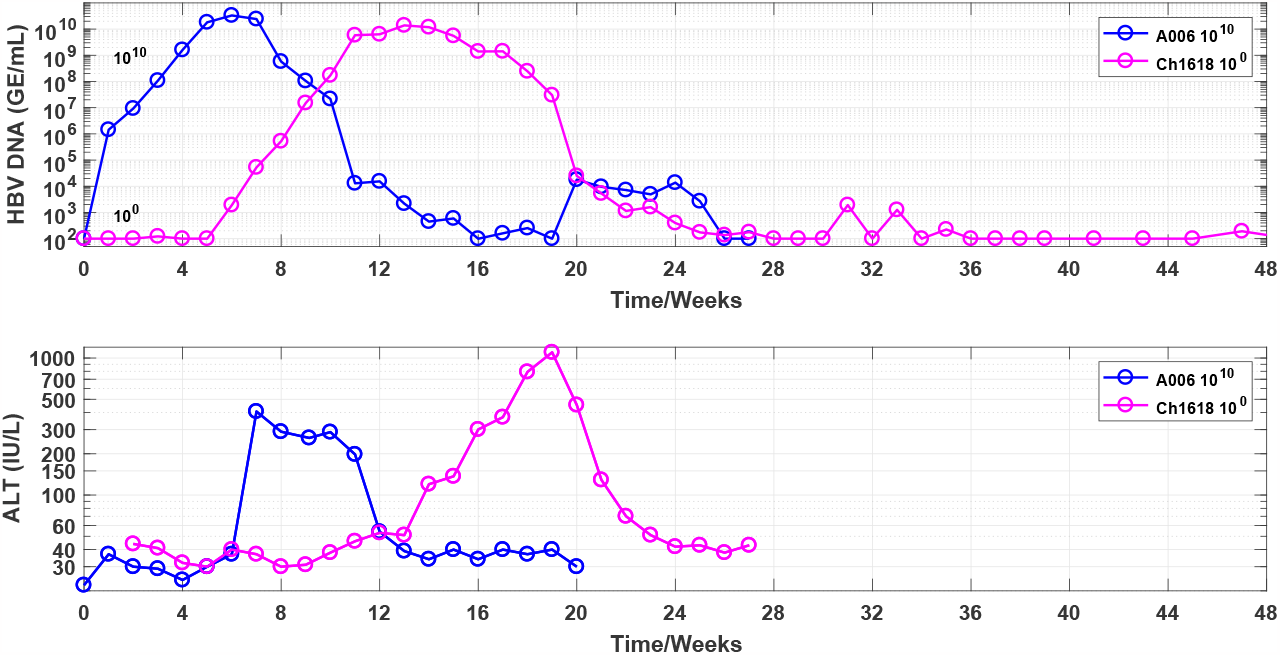
The outcomes of HBV DNA and ALT in chimpanzee A006 with 10^10^ GN/anima and chimpanzee Ch1618 with 1GN/animal (Thank Dr. S. F. Weiland at The Scripps Research Institute for proving the experimental data of the chimpanzees).

Observe that:

- The peaks of the HVB DNA of the seven chimpanzees were between 3.875× 10^7^ GN/mL (Ch007) and 3.3125× 10^10^ GN/mL (Ch006).
- After immune responses activated, the HVB DNA of five of the seven chimpanzees was lower than undetectable level 100 GE/mL (100 GE/mL was the lower limit of the detection for the assays) at or before week 41.
- The HVB DNA levels of four of the five chimpanzees have relapsed shortly, and another has relapsed longer.
- From Fig. 6, it follows:
  a. The peaks of the HVB DNA of the two chimpanzees have been higher than 10^10^ GN/mL.
  b. The peak day of the HVB DNA levels of chimpanzee Ch1618 has been later seven weeks than the peak day of the HVB DNA levels of chimpanzee Ch006.
  c. The HVB DNA levels of the two chimpanzees have relapsed shortly and longer, respectively.
  d. Before the ALT levels were higher than 40IU/L, the HVB DNA levels of the two chimpanzees had been higher than 10^8^ GN/mL for several weeks.

Theoretically, the virus infection models [9, 10] show that if one infected individual’s basic virus productive number *R*_0_ is > 1, the individuals will be obtained symptomatic or asymptomatic continued infection even if infected with one virus strain. Otherwise if their *R*_0_ *<* 1, the infected individuals will recover finally even if infected with a large amount of virus. The simulations to the dynamic of two acute HBV infected chimpanzees shows that the chimpanzee Ch1603’s activated immune ability had disappeared during the HBV DNA relapsed periods [10]. We guess that some irregular living habit and/or other sudden events (for example catching a cold) made the chimpanzees’ immune abilities have reduced temporarily, which caused their HBV DNA to relapse shortly or longer [10].

The theoretical results suggest that before their immune responses were activated, the seven chimpanzees’ basic virus productive number *R*_0_ were large than 1, after that six chimpanzees’ basic virus productive number *R*_0_ were less than 1.

The above stated experimental and theoretical results may help us to understand the phenomena appearing in COVID-19 epidemics. It can be assumed:

- Some people may never be infected by COVID-19 even infected with high dose COVID-19 virus because their basic virus productive number *R*_0_ *<* 1.
- Some people may become infected COVID-19 even infected just one COVID-19 virus if their basic virus productive number *R*_0_ *>* 1.
- Some people may be infected by the COVID-19 virus coming from the asymptomatic infected individuals who’s SARS-CoV-2 nucleic acid level were too low to be detected.

WHO has provided brief and key answers on COVID-19 and related health topics [11]. However the WHO’s technic guidelines cannot be obtained complete or timely implements by some administrative authorities and/or people in some countries. Hence some countries have experienced multiple outbreaks of COVID-19 epidemics. The paper recommends some suggestions (referring [11]) as follows:

1. In regions appearing new COVID-19 variant infection. Suggest:
  a. People should implement more strict personal prevention measures including using medical masks in gatherings, avoiding crowds and close contact, regularly cleaning hands, keeping rooms well ventilated if save distance (at least a 1-metre distance from other infected people) cannot be kept. (see [11] for detail)
  b. All administrations act quickly to discover and extinguish together, quickly cut off the transmission chain until the community transmission of COVID-19 has been initially blocked. For this purpose, the administrations should at least maintain the prevention and control measures implemented 7 days after reaching the turning point [2, 3, 5].
  c. Using more accuracy SARS-CoV-2 nucleic acid testing (CT value *>*40) discovers potential infected individuals.
  d. Individual who do not have symptoms but have had close contact with someone who is, or may be, infected may take COVID-19 antigen detection at least one time every week.
2. In regions of the numbers of current symptomatic and asymptomatic infected individuals decrease significantly.
  a. Most people’s *R*_0_ *<* 1. They can prevent presenting COVID-19 virus infection and do not need to implement special preventions to the COVID-19 virus.
  b. The administrations in the regions should implement effectively prevention and control measures to prevent and discover (same as (1) (c)) outside region input new COVID-19 variant virus infected individuals.
3. In regions only finding a few outside input new COVID-19 variant virus infected individuals. All administrations should implement the measures in (1) (b) and (c). The individuals should be implement the measures in (1) (d).
4. In regions ending old COVID-19 epidemic and not finding new COVID-19 variant virus infected individuals. The administrations should implement the measures in (2) (b). The people do not need to implement special preventions to COVID-19 epidemic.
  - In summary, if you don’t know who is infected by new COVID-19 variant virus, you should assume that your *R*_0_ *>* 1 to the new COVID-19 variant virus. Just one COVID-19 variant virus entering your mouth or nose by breathing or touching COVID-19 variant virus polluted surfaces can get sick with COVID-19 and become seriously ill or die [11].

## 6 Concluding Remarks

The main contributions of this paper are summarized as follows:

1. It is the first time to summary the COVID-19 epidemic from December 31 2021 to December 6 2022 in mainland China. It shows a clear picture to prevent and control the spread of the COVID-19 in mainland China epidemics [4].
2. It uses two models to simulate the recent mainland China epidemics. The simulation results on the end points of the investigated time intervals were in good agreement with the real word data [4], in particular the case for foreign input infections.
3. The simulation results can provide possible interpretations and estimations to the prevention and control measures, and the effectiveness of the treatments.
4. The simulations showed that the average input transmission rate of the foreign input symptomatic infection individuals was much lower than the average transmission rates of the symptomatic infection causing by the mainland symptomatic and asymptomatic individuals.
5. The simulations showed that the average input transmission rate of the foreign input asymptomatic infection individuals was was much lower than the average transmission rate of the asymptomatic infection causing by the mainland symptomatic individuals.
6. The simulations showed that the average recovery rates of the foreign input COVID-19 symptomatic and asymptomatic infected individuals were much higher than the average recovery rates of the mainland symptomatic and asymptomatic infected individuals.

For the mainland domestic epidemic visual simulations:

- If kept the transmission rates, the recovery rates, the death rate and the blocking rates on day 181 (June 30, 2022), the numbers of the current symptomatic and asymptomatic individuals would reduce to about one on day 270 (September 27, 2022).
- If kept the transmission rates, the recovery rates, the death rate and the blocking rates on day 340 (December 6, 2022) until day 380 (January 15, 2023), the numbers of the current symptomatic and the asymptomatic infected individuals would increase to 37 999 and 224 499, respectively, the cumulative death individuals would increase from 599 to 616, respectively.
- If kept the transmission rates, the recovery rates on day 340, but decreased blocking rates to 34% and select the death rate to equal to the average death rate during days 104-150, then the simulation showed that on day 380, the numbers of current symptomatic and the asymptomatic infected individuals would increase to about 337 238 057 and 501 626 885, respectively, and the cumulative death individuals would reach about 1 055 607.

For the foreign input epidemic visual simulations:

- If kept the transmission rates, the recovery rates, and the blocking rates on day 99 (April 9, 2022) until day 340, the numbers of the current symptomatic and the asymptomatic infected individuals would decrease to about 1 and 40, respectively on day 215 ((August 3), and 0 and 1 on day 340, respectively.
- If kept the transmission rates, the recovery rates, the death rate and the blocking rates on day 340 until day 380 (January 15, 2023), the numbers of the current symptomatic and the asymptomatic infected individuals would decrease and increase to 168 and 1952, respectively.
- Different combinations of the eight parameters of Equation (1) may generate similar simulation results. Therefore need further study to obtain better parameter combinations to interpret COVID-19 epidemics.

It is not wise strategy to withdraw all prevention and control measures before the number of the all social infected cases have been cleared. 100% blocking the speed at which COVID-19 infection spreads is key strategies for early clearance or reduction of epidemic spread possible [2, 3, 5, 6].

It is necessary that administrations implement strict prevent and control strategies to prevent the spread of new COVID-19 variants. It is expected that spreads of new COVID-19 variant infection can be stopped soon if the recommends are implemented.

## Data Availability

http://www.nhc.gov.cn/

## Data availability statement

Data are available on reasonable request. Please email the author.

## Ethical Statement

Not applicable/No human participants included.

## Acknowledgement

The author gratefully acknowledge Ms Aiquan Min at Peking University for checking the dataset.

